# Finding the Forest in the Trees: Using Machine Learning and Online Cognitive and Perceptual Measures to Predict Adult Autism Diagnosis

**DOI:** 10.1101/2025.03.13.25323899

**Authors:** Erik Van der Burg, Robert M. Jertberg, Hilde M. Geurts, Bhismadev Chakrabarti, Sander Begeer

**Author notes:** Corresponding author: Robert Jertberg. Both authors contributed equally to this work (Erik Van der Burg, Robert M. Jertberg).

## Abstract

Traditional subjective measures are limited in the insight they provide into underlying behavioral differences associated with autism and, accordingly, their ability to predict diagnosis. Performance-based measures offer an attractive alternative, being designed to capture neuropsychological constructs more directly and objectively. However, due to the heterogeneity of autism, differences in any one specific neuropsychological domain are inconsistently detected. Meanwhile, protracted wait times for diagnostic interviews delay access to care, highlighting the importance of developing better methods for identifying individuals likely to be autistic and understanding the associated behavioral differences. We disseminated a battery of online tasks measuring multisensory perception, emotion recognition, and executive function to a large group of autistic and non-autistic adults. We then used machine learning to classify participants and reveal which factors from the resulting dataset were most predictive of diagnosis. Not only were these measures able to predict autism in a late-diagnosed population known to be particularly difficult to identify, their combination with the most popular screening questionnaire enhanced its predictive accuracy (reaching 92% together). This indicates that performance-based measures may be a promising means of predicting autism, providing complementary information to existing screening questionnaires. Many variables in which significant group differences were not detected had predictive value in combination, suggesting complex latent relationships associated with autism. Machine learning’s ability to harness these connections and pinpoint the most crucial features for prediction could allow optimization of a screening tool that offers a unique marriage of predictive accuracy and accessibility.

---

Autism is associated with a uniquely pervasive collection of behavioral differences, manifesting at multiple stages from basic sensation to fine motor skills to complex cognitive processes ^1–5^. On an individual level, the degree to which each related area may differ from the norms seen in the non-autistic population is highly variable, which has translated into inconsistencies in research outcomes and challenges for clinicians in diagnosing the condition. While there are certain areas, such as multisensory perception ^2^, emotion recognition ^4^, and executive function ^1^, where meta-analyses show especially pronounced differences between autistic and non-autistic individuals^1^, they still note considerable heterogeneity (exacerbated by small samples), and null findings are common in the literature. What is more, the effect sizes of studies comparing individuals with and without autism across a range of neurocognitive constructs have declined in a linear trend over the past two decades, from 45-80% in some domains ^7^. This suggests that objective measures of behavioral differences may not have the same ability to distinguish between autistic and non-autistic individuals that they did in the past. Despite this, they have clear importance both to theoretical understanding of the underlying neuropsychological differences that characterize the condition and to the development of tools that could identify individuals likely to be autistic in a manner that circumvents biases/inaccuracy in the assessment of others or self-report ^8–13^.

Additionally, although adults comprise the vast majority of individuals with autism, research focuses primarily on children ^14,15^. While this limits our understanding of autism throughout the lifespan, cross-sectional research and meta-analyses suggest that some differences between autistic and non-autistic individuals may diminish in magnitude or even be resolved entirely by adulthood ^1,2,16–18^. Our recent research with a large sample of adults with and without autism conforms to this trend. While we detected behavioral differences in a range of online tasks measuring multisensory perception ^19^, emotion recognition ^20^, and executive function ^21^, we also failed to replicate numerous findings from earlier research involving smaller samples, primarily of children. Most of the significant differences we detected were in secondary variables and novel analyses investigating the temporal features of performance, highlighting the importance of developing new methods to gain insight into objective behavioral differences between autistic and non-autistic individuals.

This endeavor is of unique importance among autistic adults for several reasons. Many develop masking behaviors that might render subjective insight into their condition less reliable ^22–24^. Additionally, those diagnosed later tend to score lower on questionnaires designed to capture autistic traits ^25^. In fact, among adults, the most common questionnaires used in research, screening, and assessment are quite lackluster predictors of diagnosis ^26–29^ and may be especially subject to report ^13^ and gender biases ^8^, as well as cross-cultural differences^30,31^. This predictive imprecision may be due in part to the fact that many were designed primarily with children in mind. These findings suggest that the traditional subjective avenues of diagnostic assessment and self-report might more frequently fail to distinguish between autistic and non-autistic adults than children and may suffer from certain biases that should not affect more objective measures of performance. Given that the rate of adult diagnoses increased twentyfold and mean age of diagnosis rose from 9.6 to 14.5 years between 1998 and 2018 according to data from the United Kingdom ^32^, the need for more sensitive objective measures of behavioral differences to supplement subjective ones is increasingly urgent, in contrast with their apparently diminishing ability to discriminate between groups. These trends in rates of diagnosis may be due in large part to increasing demand for diagnostic assessment leading to highly protracted wait times, further highlighting the utility of inexpensive, easily accessible tools that might indicate those most likely to be autistic for expedited assessment and support.

One avenue for pursuing this objective is to look beyond specific, independent behavioral measures and towards collections of differences that span across the related domains of function. It is possible that focusing on the former could risk missing the forest for the trees and that trading small, inconsistently observed individual effects for more robust profiles of differences could greatly enhance sensitivity to autism. Multivariate statistical approaches can turn modest individual effects into large combined estimates of group differences ^33^, but lie somewhere between the average and sum of these individual components, depending on the strength of their correlations. Machine learning techniques are ideally suited for making more of large datasets, creating a model that is greater than the sum of its combined parts. They are able to capture complex, multidimensional relationships among variables that give them greater meaning in the context of other data than they have in isolation. One application of this approach is in predicting the classification of a certain data type (such as whether or not an individual has a diagnosis) given performance on a range of tasks. Tree ensemble methods like random forests (RF) are popular tools for processing high-dimensional datasets in this manner due to their ability to detect sparse signals, resulting in good predictive accuracy ^34^. They capture any patterns that exist in the data, and are therefore difference focused rather than deficit focused. Additionally, RFs provide insights into feature importance, ranking features (i.e., variables) by their contribution to the model. This ranking is valuable for feature selection and understanding the data structure underlying prediction. It could thereby be informative in selecting the measures most sensitive to autism in a battery of tasks and elucidating the profile of behavioral differences that characterizes the autism spectrum.

Previous studies have demonstrated the effectiveness of machine learning approaches to predicting autism, primarily using neurological biomarkers ^35–38^. However, neuroimaging techniques are costly (particularly the frequently employed magnetic resonance imaging), time-consuming, and require participants to be present in the lab. Subjective measures like questionnaires offer an alternative that is more accessible and affordable, and combinations of parental reports about children and machine learning have demonstrated good predictive accuracy ^39^. However, research with adults is more limited and is still subject to the aforementioned biases. Online, performance-based measures offer a marriage of more objective data types with the accessibility of surveys. A recent study combining performance-based data types and parental report with machine learning was able to demonstrate high predictive accuracy for autistic children ^40^. However, no study to date has attempted to use a combination of online tasks and machine learning to predict autism diagnosis for adults, who have proven the most difficult to distinguish using traditional assessment tools. Our large dataset, spanning across behavioral domains relevant to autism, makes us uniquely well positioned to investigate whether such a battery of tasks can predict autism effectively and to use feature ranking to select the most useful measures in doing so. This may not only offer insight into which variables/measures are important for diagnosing adults, it could also allow future expansion and fine-tuning of such a battery to optimal predictive performance. To this end, we employed an RF algorithm to predict whether participants were autistic or not using 54 distinct variables we collected in our series of experiments (see the Supplementary Materials for details). We then compared this performance to that of the most popular autism screening questionnaire for adults, the Autism Quotient (AQ) 28 ^41^, and measured how much combining data types could improve performance.

## Methods

### Dataset

We reused data from a series of experiments measuring multisensory perception, emotion recognition, and executive function using overlapping participants ^19–21^. Initially, 538 Autistic participants were recruited via the Netherlands Autism Register (NAR, https://nar.vu.nl/), and 421 non-autistic participants were recruited via Prolific Academic as well as the NAR. Autistic participants reported having a formal diagnosis by a registered clinician; non-autistic participants reported no autism diagnosis. All participants were fluent in Dutch, naïve to the purpose of the experiments, and provided informed consent prior to the experiments. The studies were approved by the ethical committee from the Vrije Universiteit Amsterdam (VCWE-2020-041R1) in accordance with the Netherlands Code of Conduct for Research Integrity and the revised declaration of Helsinki. All experiments were programmed and conducted online using Neurotask (www.neurotask.com).

A total of 407 participants were excluded as data from at least one of the experiments was missing (due to exclusion criteria as mentioned in each study, technical issues, or attrition). As a result, the final dataset consisted of 286 autistic individuals and 266 non-autistic individuals. For the 100% age- and gender-matched group (n=125 per group), we randomly selected individuals from our unbalanced sample. Table 1 depicts the demographic information for the participants included in this study.

**Table 1.** Demographic information over all participants and for the age- and gender-matched group. Asterisks signify significant group differences (*p* < .001), as revealed by Mann-Whitney U tests.

|  | All participants |  |  |  | Age/gender-matched participants |  |  |  |
| --- | --- | --- | --- | --- | --- | --- | --- | --- |
|  | Autistic |  | Non-autistic |  | Autistic |  | Non-autistic |  |
|  | (N = 286) |  | (N = 266) |  | (N = 125) |  | (N = 125) |  |
| Age: mean (SD) | 44.2 | (13.6) | 32.2 | (12.0)* | 37.6 | (12.7) | 37.6 | (12.7) |
| Gender: m/w (%w) | 203/83 | (71.0) | 129/137 | (48.9)* | 93/32 | (74.4) | 93/32 | (74.4) |
| ICAR: mean (SD) | .54 | (.20) | .53 | (.17) | .54 | (.21) | .55 | (.19) |
| AQ total: mean (SD) | 83.4 | (10.7) | 60.1 | (11.6)* | 81.6 | (11.0) | 59.7 | (12.3)* |

AQ: Autism Quotient <sup>42</sup>.
ICAR: International Cognitive Ability Resource (abbreviated intelligence quotient test) <sup>2</sup>

We extracted 54 key features from three studies. In the first ^19^, we employed the McGurk/MacDonald paradigm (mcg in Figures 2 and 3) to assess participants’ ability to integrate audiovisual speech stimuli ^46^, to process multisensory temporal information ^47^, and to rapidly adapt to multisensory asynchronies ^48–50^. In the second ^20^, we evaluated performance on three emotion recognition tasks: basic facial emotion recognition (kdf) ^51^, complex facial emotion recognition (rme) ^52^, and affective prosody recognition ^90^. In the third ^21^, we assessed four executive functioning tasks: response inhibition (gng) ^54^, visual orienting (agc) ^55^, spatial working memory (cbd) ^56^, and cognitive flexibility (tmt) ^57^. See the Supplementary Materials for details regarding each feature and a brief description of each task. Furthermore, we included two additional features representing age and gender, making a total of 56 input features. This was done to get an accurate representation of the features important to predicting autism in the overall sample. Without age/gender included as factors, given disparities between groups in these factors, the feature ranking might reflect variables that are good predictors of age/gender rather than (or in addition to) diagnosis. While including these factors inflated performance in the overall sample, it had no influence on the matched subsample. The data were not normalized as decision trees like RF are typically not affected by the scale of the input features.

### Data preprocessing

The machine learning algorithm was programmed using Python (version 3.11.7), and the code can be found at https://osf.io/nxyzt/?view_only=b9b60eeaf0f54495aa81fb2fc4955d97. First, we fixed the random seed (1974) to make it feasible to reproduce the outcomes of the classifiers. Then, the train_test_split function of the Scikit-Learn module was applied to split the dataset into a training (80 %) and test dataset (20 %). Provided that both groups were rather equally represented in our training dataset (i.e., 50.3 % versus 49.7 % for individuals with and without autism, respectively), there was no reason to suspect that the model was biased towards the majority class. Therefore, we decided not to oversample the minority class using e.g. synthetic minority over-sampling technique (SMOTE ^58^) or, alternatively, to exclude participants from the majority class to balance the datasets.

### Model optimization and evaluation

The hyperparameters were tuned to optimize the RF model and to avoid overfitting through 3-fold cross validation using the GridSearchCV function implemented in Scikit-Learn module. Subsequently, we set the hyperparameters to the optimal parameters determined by the GridSearchCV function, and systematically manipulated each parameter to visualize the impact on the model performance (i.e., F1 score). Overfitting was avoided by adjusting parameters to minimize performance differences between training and test datasets (as a large disparity between training/test performance is indicative of overfitting). The hyperparameters were *n_estimators* (number of trees in the forest), *max features* (max number of features considered for splitting a node), *bootstrap* (method for sampling data points), *max_depth* (max number of levels in each decision tree), and *min_samples_leaf* (min number of data points allowed in a leaf node). This procedure was performed for both datasets (i.e., all participants included and the age/gender-matched participants). When all the participants were included, the hyperparameters were set to: n_estimators = 100, max_features = ‘sqrt’, bootstrap = True, max_depth = 4, and minimal_samples_leaf = 60. When only the age/gender-matched participants were included, the hyperparameters were set to: n_estimators = 100, max_features = ‘sqrt’, bootstrap = False, max_depth = 3, and minimal_samples_leaf = 60.

The performance of a model was derived by measuring the number of true positives (TP; i.e., the number of autistic individuals who were correctly classified as autistic), true negatives (TN; i.e., the number of non-autistic individuals who were correctly classified as non-autistic), false positives (FP; i.e., the number of non-autistic individuals who were incorrectly classified as autistic) and false negatives (FN; i.e., the number of autistic individuals who were incorrectly classified as non-autistic). We used these to calculate each model’s accuracy: (TP+TN)/(TP+FP+FN+TN), sensitivity: TP / (TP + FN), specificity: TN / (TN + FP), and F1 score: 2*TP / (2*TP+FP+FN). The sensitivity signifies a model’s ability to correctly classify autistic individuals as being autistic, whereas the specificity signifies the models ability to correctly classify non-autistic individuals as being non-autistic. The F1 score reflects the harmonic mean of sensitivity and specificity, which is a better metric of incorrectly classified classes than the accuracy. As such, F1 scores are a better reflection of the balance between sensitivity and specificity. However, for transparency and comparability with other studies, all values are reported.

The RF model performed this classification procedure 1000 times, and we extracted the feature importance from each iteration to understand what features drive the predictions by averaging their importance over all models. To determine the significance of these features, we reran the model 1000 times using a random RF model. This model used the same dataset and identical hyperparameters but with randomized autism labels, effectively creating a control group. The feature importance extracted from these random RF models was averaged to serve as a baseline (see these studies ^59–61^ for a similar approach). Finally, for each feature, we conducted a two-tailed independent *t*-test to examine whether the feature importance for the RF model was significantly larger than the feature in importance for the random RF model (α was set to .05, and *p* values were Bonferroni corrected). Note that all steps were performed twice (once with all participants and once with only the age/gender-matched participants).

## Results

### Model performance

Figure 1 (right panels) illustrates the mean performance (accuracy, sensitivity, specificity, and F1 score) for the RF classifier model (orange bars) and the random RF classifier model, where group labels were randomized during training to create a baseline comparison (gray bars). Whereas the dark bars signify the performance on the training dataset, the light bars reflect the performance on the test dataset. For the upper panel, all participants (286 autistic individuals and 266 non-autistic individuals) were included in the model. Note that the autistic individuals were significantly older than the non-autistic individuals (see the age distribution in Figure 1), and that the ratio of men to women differed significantly between the two groups. Given that age as well as gender can have a profound effect on multisensory processing ^62,63^, emotion recognition ^64,65^, and executive function ^66^, we decided to rerun the RF model on a 100% age- and gender-matched group (N = 125 per group) as well. For the age/gender-matched participants, the performance of the RF and random RF classifier are shown in the lower panel.

**Figure 1.**
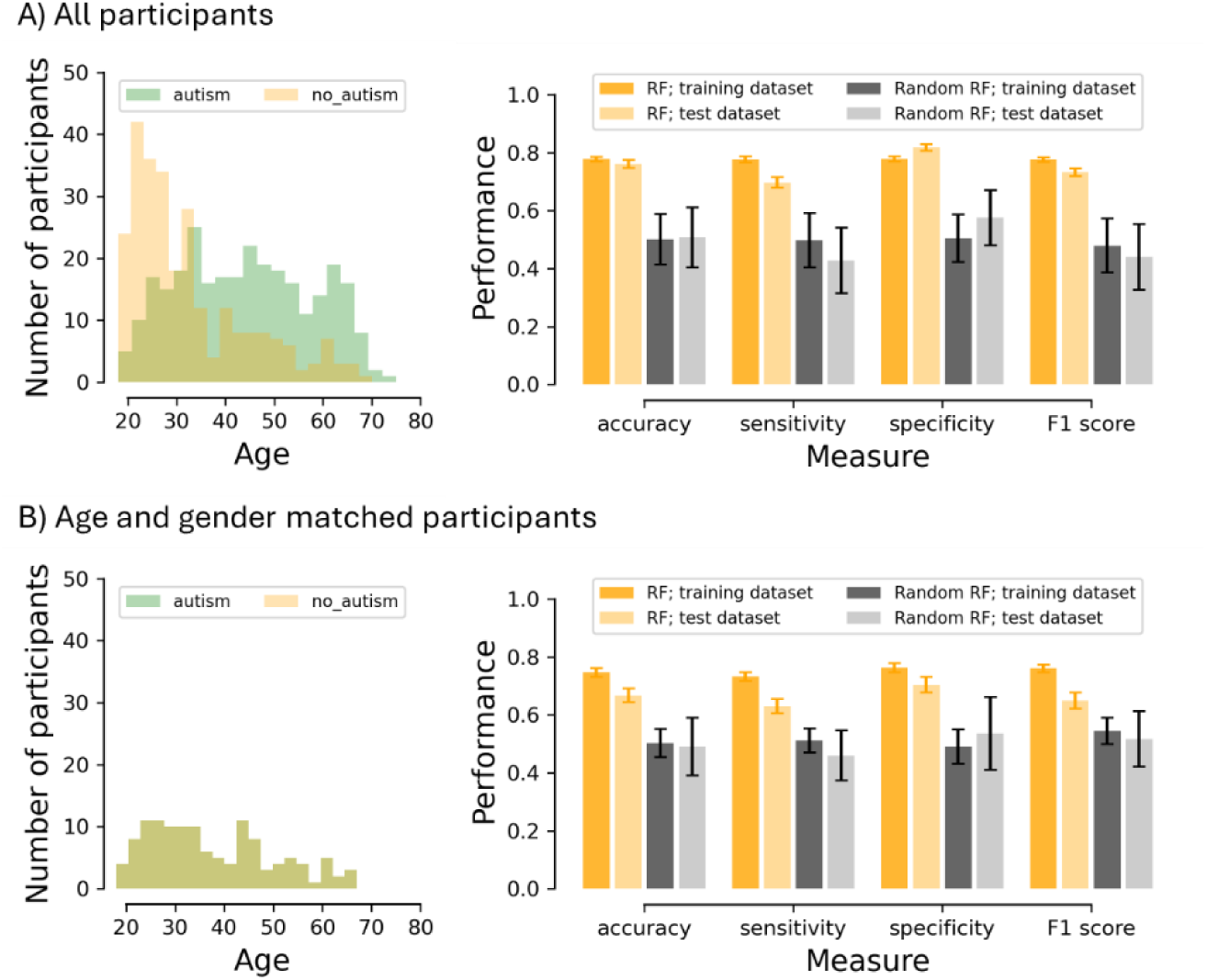
Age distribution and model performance for all participants (panel A) and for age- and gender-matched participants (panel B). Left panels reflect the age distributions. Right panels reflect the mean performance for both the random forest (RF) classifier models with properly labeled groups (orange bars) and the RF classifier models with the group labels randomized (gray bars) in terms of accuracy, sensitivity, specificity and F1 score. The dark bars signify the performance on the training dataset, whereas the light bars reflect the performance on the test dataset. Error bars represent the standard deviation.

As is clear from Figure 1 (right panels), for the RF model (orange bars), we were able to predict autism both among all participants and the age- and gender-matched subsample with reasonable accuracy. As expected, when we randomized the labels during training (i.e., the random RF model), performance was close to chance level when we tested performance on the original training and test dataset (gray bars). In contrast, when fed the correct labels, our best random forest model (i.e., the one with the highest F1 score) achieved a test accuracy of 81.8%, sensitivity of 77.1%, specificity of 84.1%, and F1 of 77.9% for the full sample. For the age/gender-matched sample, these values were 74.0%, 69.2%, 79.2%, and 73.5%, respectively. Note that performance in the unmatched sample is inflated by the inclusion of age/gender as features, given the disparity between groups in these factors. As such, the matched sample is a better estimate of performance (although it is limited by the smaller sample size).

### Feature importance

Feature importance is a measure that assigns a score to each input feature of the dataset, indicating its relevance in predicting autism. Figures 2 and 3 illustrate the mean feature importance score for each feature as well as the likelihood that each of the others will appear in a decision tree with them for the full and age/gender-matched datasets (respectively). Here, darker blocks indicate features that frequently co-occur and are, therefore, useful in combination for predicting autism. Decision trees reflect the underlying structures by which individuals are classified according to thresholds in various features. Note that, in Figures 2a and 3a, each task was signified by a unique color. Important features are indicated by an asterisk, meaning that the feature score for the RF model was significantly larger than the feature score for the random RF model (see the dashed line). Supplementary Figures 1 and 2 contain the partial dependence plots for each significant feature for all participants and the age/gender-matched group, respectively. A partial dependence plot visualizes the functional relationship between specific features and model predictions while keeping other variables constant to extract meaningful insights.

**Figure 2.**
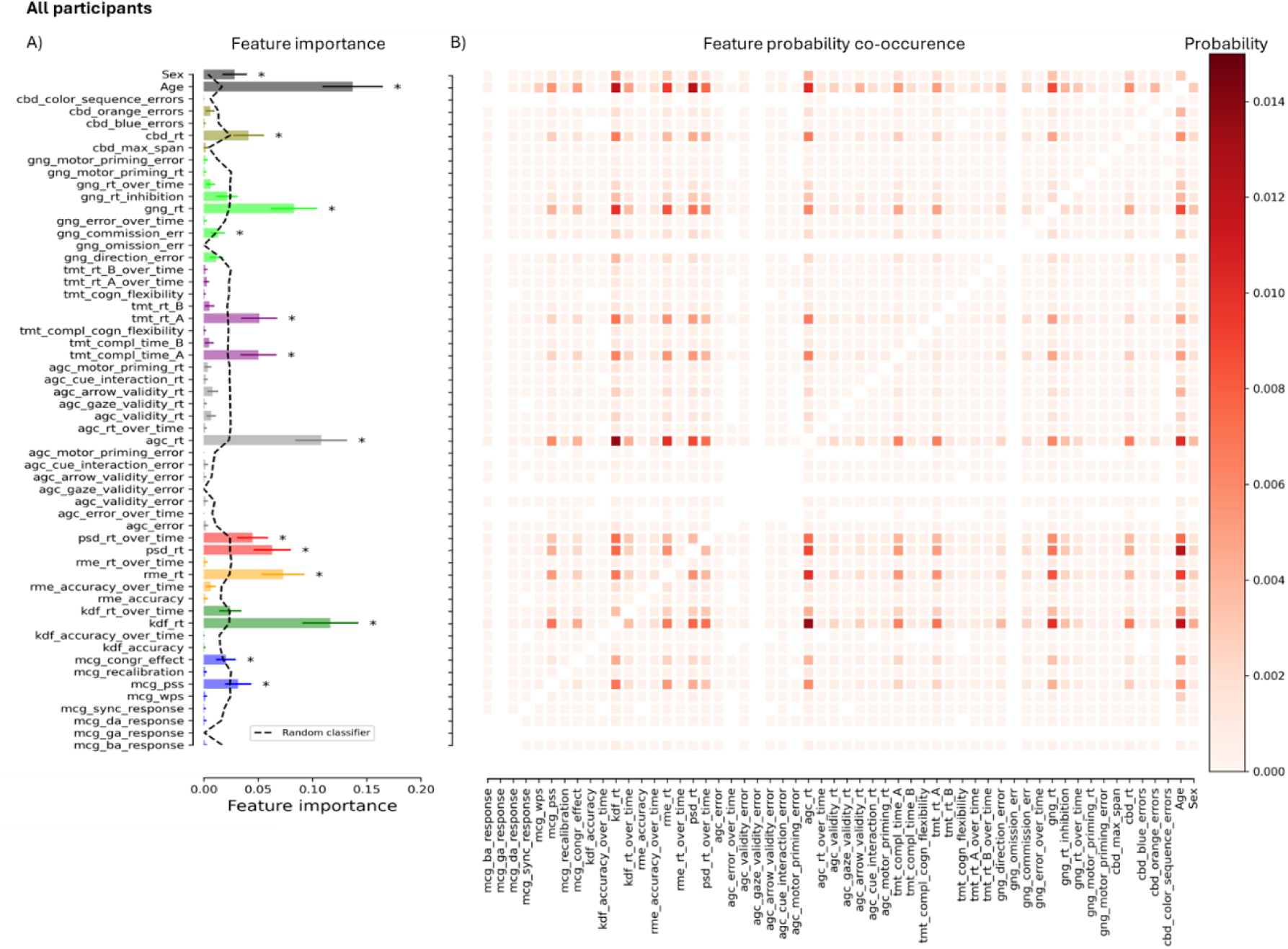
A) Mean feature importance score for each feature in the full dataset. Note that each task is identified by a unique color and the first three characters of the corresponding feature names (mentioned in the Methods section). Here, important features are indicated by an asterisk, indicating that the feature score for the RF model was significantly (*p* < .05, Bonferroni corrected) larger than the feature score for the random RF model (see the dashed line). Error bars represent the standard deviation. B) Likelihood that features will occur together in the same decision tree (all values add up to one). Darker blocks indicate features that frequently co-occur and are, therefore, useful in combination for predicting autism.

**Figure 3.**
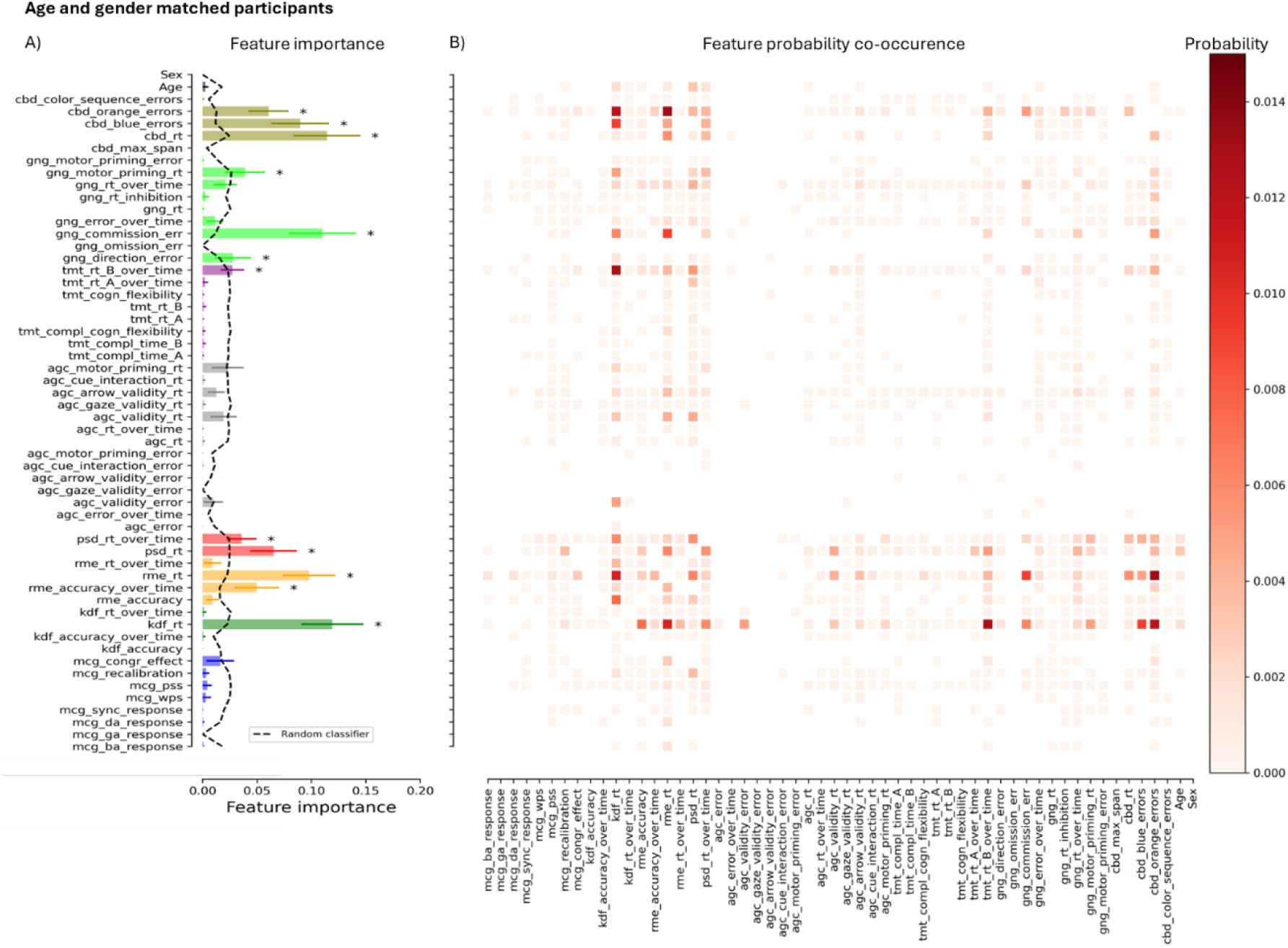
A) Mean feature importance score for each feature in the age- and gender-matched dataset. Note that each task is identified by a unique color and the first three characters of the corresponding feature names (mentioned in the Methods section). Here, important features are indicated by an asterisk, indicating that the feature score for the RF model was significantly (*p* < .05, Bonferroni corrected) larger than the feature score for the random RF model (see the dashed line). Error bars represent the standard deviation. B) Likelihood that features will occur together in the same decision tree (all values add up to one). Darker blocks indicate features that frequently co-occur and are therefore useful in combination to predicting autism.

For both the full sample and age/gender-matched subgroup, most of the important features to the model were those related to RTs, particularly on the emotion recognition tasks. However, the samples differed in that several EF measures that were not significantly more predictive than in the random RF model for all participants were in the age/gender-matched sample, whereas other EF measures were not. Fluctuations in accuracy over time in the complex facial emotion recognition task (rme) also reached significance in the matched sample alone, whereas multisensory perception variables only held importance in the full sample. While age and gender were, naturally, important features when the groups were unbalanced on these factors, they ceased to hold significance in the matched group. Please note that differences in feature importance between the samples may be due to either the influence of age/gender related factors or differences in the properties of the RF models (seen in the hyperparameters described in the Methods section). With the greater volume of input provided by the full sample, more complex decision structures were possible (reflected, for example, in the greater depth of the trees), which can also influence the features that gain importance. Features were found to co-occur with each other in distinct patterns (in contrast to Supplementary Figures 3 and 4 depicting feature co-occurrence when labels were randomized) for both the full and matched subgroups. However, these appeared to be more specific in the latter.

### Combination and comparison with the AQ-28

To determine whether our battery of online tasks could improve upon the predictive performance of the AQ-28, we added the total score from the questionnaire as an additional feature in the model (without adjusting hyperparameters so that any improvement of the model cannot be assigned to model differences) and evaluated performance for the age- and gender-matched group. Figure 4 juxtaposes the performance metrics of the best resulting RF model (see dashed lines) with those of the AQ-28 alone (see solid lines). Here, the performance is plotted as a function of the AQ thresholds (i.e., individuals were classified as autistic if their AQ total score ≥ AQ threshold).

**Figure 4.**
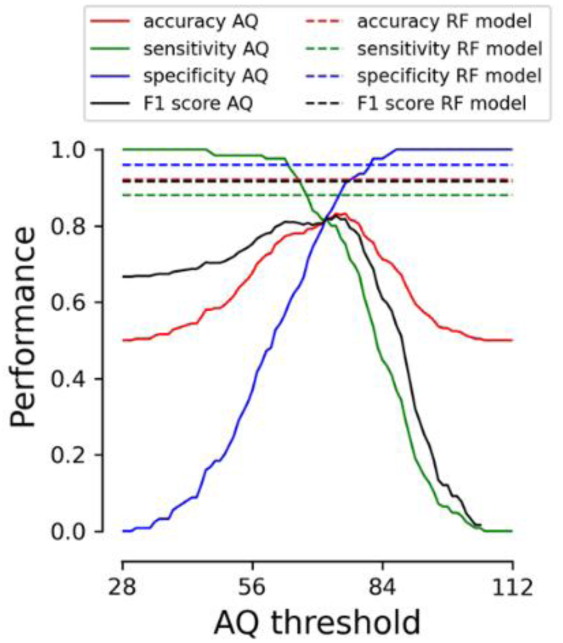
Performance metrics of the best random forest model including the objective measures and Autism Quotient (AQ-28) compared to those of the AQ-28 alone (i.e., individuals were classified as autistic if their AQ total score ≥ AQ threshold). All values correspond to the age- and gender-matched group.

The maximum accuracy of the AQ-28 within our age- and gender-matched group was 83% with an AQ threshold of 74 (i.e., all of those scoring below 74 were classified as non-autistic). In comparison, the best combined RF model was able to predict diagnosis with 92% accuracy. Supplementary Figures 5 and 6 illustrate the mean performance of the model when we included the AQ-28 and the feature importance when the AQ-28 was excluded (left panel) or included (right panel) for the age/gender-matched participants, respectively.

## Discussion

Our series of prior experiments conformed to the general trend of diminishing effect sizes and inconsistent reproducibility in the autism literature^7^, with many primary dependent variables not differing in terms of group means. Despite this, looking beyond the individual measures and towards differences between groups in the broader behavioral profile provided a promising start for the marriage of machine learning and online cognitive/perceptual testing. Even if the subtle differences in the timbre of each instrument might not be noticeable, the tune of the music they offered in concert was of recognizably different character to our machine learning algorithm. It was able to distinguish between groups quite well using only data from our novel battery of objective measures, and combining them with the most popular autism screening questionnaire for adults led to a significant improvement upon the predictive accuracy of either data type alone. The RF (random forests) model was also able to reveal the manner in which specific features gained value in combination and contributed to this impressive performance, allowing insight into the underlying profile of neuropsychological differences that characterizes autism. While there are, of course, limitations to the conclusions that can be drawn from these initial efforts, there are also many opportunities for improvement, particularly in how what we have learned on the featural level can inform enhancement of performance on the holistic level.

On the former level, in both our full sample and age/gender-matched subsample, variables where significant differences between groups were detected consistently (particularly those involving RT) were, unsurprisingly, of high predictive value. More intriguingly, certain variables where significant differences were not detected in group means were found to be of value in combination in predicting autism diagnosis, even after highly conservative Bonferroni corrections. These include variables related to the time taken to complete the two parts of the trail making task (which is used to measure cognitive flexibility but is also influenced by visual search and motor behavior), the rate of errors of commission on the go/no-go task (indicative of failures in inhibition), a secondary category of errors on the chessboard task (suggesting working memory issues), and features reflecting subtler effects like fluctuations in performance over time. It is unclear whether the RT differences are due to domain specific processing differences or generalized differences in factors like motor speed and meta-cognition. However, most of these other variables are related to executive function, an area in which effect sizes derived from comparing group means have declined significantly over the years ^7^ despite their centrality to early theories of autism ^67–70^ and the prevalence of issues therein reported by autistic individuals ^71,72^. The fact that variables that did not differ significantly between groups were useful in combination in predicting autism suggests differences between groups that are embedded in latent relationships among neuropsychological features, which cannot be detected by the standard practice of comparing individual means.

These relationships can be visualized in the feature co-occurrence probability graphs (Figures 2b and 3b). In contrast to the ones for the RF models predicting the randomized group assignment (Supplementary Figures 3 and 4), there are distinct patterns in the frequency with which these variables appear together in decision trees, reflecting a logic to the manner in which they are used to classify individuals. It is important to note that, despite superficial resemblance, these are not correlational matrices; they simply reveal the probability that a decision tree including one variable also includes another. This could mean that, for example, individuals who show differences in feature a *and* feature b are more likely to be autistic, or that individuals who show differences in feature a *or* feature b are more likely to be autistic. As such, the frequent co-occurrence of variables does not imply a direct correlation between them. In fact, if variables show a very strong correlation, the information they provide to the model becomes redundant, so they are less useful in combination (as almost all the individuals above/below a certain threshold in feature a will also be above/below a certain threshold in feature b).

Because of this, theoretical interpretations of the co-occurrence of features would require further investigation of the nature of their relationships. However, the figures do elucidate how different variables gain predictive value in combination (and thus the connections that must be preserved for the model to function well as a whole). For example, while the rates of commission errors do not differ significantly between groups (and would therefore be poor predictors on their own), they are ranked as important features in both the full and age/gender-matched samples (with particular significance in the latter). This is because they gain sensitivity when combined with certain other measures, most notably the reaction times on the two facial emotion recognition tasks and the rate of a certain type of errors on the working memory task. It could be that those who show differences in commission error rates and one or more of these other variables are classified as autistic, or that differences in either variable lead to autistic classification (although this is less likely for variables like commission error rates, where the groups do not differ significantly in means). In the former case, overlapping differences may define the group. In the latter, there could be subgroups of autistic individuals showing distinct behavioral differences. As such, studying the co-occurrence of features detected by the RF model can both inform us about the constellations of features necessary to improve holistic performance and direct future research towards the nature of the relationships among objective behavioral differences in autism.

Focusing on the specific measures that rank highly in feature importance and co-occur with a wide range of other prominent features could allow improvement of such a task battery. In line with our previous individual studies ^20,21^, those of other researchers ^73^, and two meta-analyses ^74,75^, these often appear to be RT variables (particularly in emotion recognition tasks) that are typically considered secondary to accuracy/error rates. Designs that allow investigation of the time course of emotion recognition could therefore be of particular value, especially ones that accentuate differences in RT. For example, certain emotional categories and types of stimuli ^76–78^ have been found to be more difficult to categorize, potentially translating into more room for RT differences. This was seen in our basic emotion recognition experiment (features coded as “kdf” in the figures), where the differences between groups in RT varied significantly according to emotional category ^20^. In this manner, what is gleaned from feature ranking can be used to fine-tune a battery by implementing manipulations that enhance insight into useful dependent variables from the most sensitive domains. This approach could also be used to drastically reduce the length of the current battery by removing the measures (or conditions) that did not generate as useful features.

It is worth keeping in mind that the most sensitive measures may differ according to factors like the age of participants, so the ranking reported in this study may not generalize to all autistic populations. In our adult sample, executive function and emotion recognition differences were of greater predictive value than multisensory ones. This is not surprising, given that multisensory differences have been found to be much more pronounced among children ^2,16,79^, whereas emotion recognition differences may be greater among adults ^4,80,81^. As such, including age as a factor and comparing the features that are important to predicting autism among children vs. adults could also allow greater insight into the most sensitive measures across development. That being said, the fact that age was not a significant feature in our matched group and did not frequently co-occur with others reveals that it was not interacting with them. This implies that the relevant features may not differ as a function of age among adults.

On the holistic level, even with objective measures alone, our model was able to predict autism quite well in both the unmatched (maximum accuracy: 82%) and matched samples (maximum accuracy: 74%). While some studies combining neuroimaging techniques and machine learning approaches have reported greater predictive accuracy than our objective measures ^38^, they are subject to prohibitive cost and time requirements relative to online testing. Questionnaires are not subject to this limitation, and some studies combining them with machine learning approaches have reported higher predictive accuracy for adults ^82–86^. However, these used the same dataset of individual AQ-10 items to predict whether participants fell above or below a certain AQ cutoff score (rather than to predict whether or not they received an official diagnosis) or used groups that were not balanced on demographic features they included as features. This risks sampling biases inflating performance, which we accounted for in our age/gender-matched analysis. It is also worth noting that in one of these studies, accuracy declined from 98% (with children) to 81% (with adults) ^83^. This is a testament to how much more difficult it is to predict autism among adults using traditional questionnaires, even with the addition of machine learning techniques. Among adults, there are reasons to suspect that the type of participants who took part in our studies would be particularly difficult to distinguish from non-autistic individuals, regardless of the methods employed (which is supported by the fact that higher AQ predictive accuracy has been reported with other samples ^87^). Our participants have average to above average IQ levels (non-significantly different from our control group), generally low support needs, and a late mean age of diagnosis ^88^. With this in mind, classification performance with our objective measures alone was a promising start.

The efforts to predict autism with machine learning described so far use one data type, be it neuroimaging data, questionnaire data, or (in our initial analyses) task performance. Yet, the approach benefits greatly from diversity in data types, where relationships among dissimilar variables can be exploited. This was seen in our combination of the objective measures with the AQ, where predictive accuracy rose to 92% (with a very similar feature importance profile to that of the objective measures alone, as seen in Supplementary Figure 6, suggesting that they offer information that is useful in a different way to the survey questions). This is a substantial improvement upon both subjective (83% max accuracy for the matched sample) and objective measures (74% max accuracy for the matched sample) alone in our dataset that even exceeds many efforts combining machine learning and neuroimaging techniques. A 9% improvement in accuracy above the most popular screening questionnaire for adults is quite significant when one considers that the global prevalence of autism is estimated to be around 1% ^89^, with recent estimates from the Center for Disease Control approaching 3% ^90^. At such scales, advancements in predictive accuracy achieved through combining diverse data types have the potential to positively impact millions of individuals.

There is also likely greater room for improvement above existing screening questionnaires among referred samples than in case/control comparisons such as this initial demonstration. Research suggests that autism screening questionnaires perform significantly worse among referred samples due to differences in prevalence rates and the fact that those who do not ultimately receive an autism diagnosis still report higher autistic traits than non-referred controls ^28^. In one recent study, it was even found that there was no significant difference at all in AQ scores between referred adults who did and did not receive an autism diagnosis, although they did show differences in performance on a task measuring emotion recognition ^91^. This suggests that objective measures may have particular value in real-world clinical settings and underscores the importance of testing a battery such as ours in a referred sample. It is likely that it would also have less predictive accuracy here, although the findings that an emotion recognition task (which generated some of our most predictive features) still detected differences is encouraging, and there is clearly more ground to be gained upon questionnaires. However, in our study, the AQ was still more sensitive than objective measures, and it serves a role that should be reinforced (rather than replaced) by them. The strong performance we achieved was undergirded by the complementary features of the two forms of data. The objective measures we employed are not known to be subject to the biases^8,13^ that affect the AQ and demonstrated high specificity, whereas screening questionnaires optimize sensitivity and are much briefer to administer. As such, their combination balances both performance-related factors and practical, clinical ones.

While our model combining objective and subjective measures is already highly accurate, it could be optimized further. The tasks in the battery were chosen to study specific underlying neuropsychological constructs, not to maximize predictive accuracy. The battery could be streamlined by eliminating those that did not contribute much to overall performance, and its accuracy could be improved by introducing more sensitive measures in their place (motor tasks and eye-tracking may be promising options that can be administered online ^3,92–94)^. The feature ranking and co-occurrence analyses demonstrated in this paper could be used to test different configurations and build an optimally efficient battery that balances time-constraints with predictive accuracy on a scale adaptive to the demands of specific applications. Additionally, we did not tune hyperparameters for the model combining subjective and objective measures, to allow comparability to that with objective measures alone. Doing so would allow improvement of the model’s performance over that which we report. Finally, RF were chosen as they are ideally suited to our tandem goals of classification and insight into the features most important to this process. However, many alternative machine learning algorithms have been applied to predicting autism diagnosis ^95–100^, and RF are not always the most accurate. As such, there is room for improvement on both the task selection and algorithmic fronts.

Although these considerations offer much room for optimism moving forward, we must be circumspect in the conclusions drawn from this initial demonstration. One limitation of our study is that our sample was quite specific and (in the case of the age/gender-matched group) smaller than ideal (which can be seen in the fact that more complex decision structures were possible with the full sample). It was comprised entirely of Dutch individuals, and our autistic group was biased towards women and those with higher IQs relative to the broader autism population. As such, before such a battery can be useful in screening for autism, further research must be conducted on a larger scale with a more diverse cohort. Fortunately, the online format makes such an endeavor feasible, and it would be valuable beyond testing performance metrics of the battery. If feature ranking were found to differ across groups, it would suggest that these autistic communities exhibit distinct underlying neuropsychological traits. In this manner, enhancing diversity is essential to assessing generalizability, and comparisons across groups could shed light upon the heterogeneity of autism presentation, which is widely suspected to be the source of inconsistent findings throughout the literature.

In conclusion, this study has demonstrated the power of combining multifarious, complementary data types collected entirely online with machine learning in predicting autism diagnosis and understanding its underlying behavioral features. By integrating objective and subjective measures, we were able to achieve a high degree of accuracy, despite working with a sample that is known to be difficult to distinguish. Moreover, we were able to offer insight into the relationships among features that made this prediction possible, unpacking the black box. On a theoretical level, the predictive value of features that were not found to differ significantly between groups in previous behavioral experiments indicates that, despite waning effect sizes in some domains, complex latent structures involving them may still consistently differ between those with and without autism. On a practical level, feature ranking also orients clinicians and researchers alike to the measures that might be most sensitive to autism and the dependent variables that designs should focus on to better capture these differences. The room for improvement with optimization of the battery and fine-tuning of the algorithm offers a viable path towards a useful tool to support a clinician. Online, performance-based testing could never replace in-person assessment, which has benefits far beyond classification. However, our findings suggest that it could provide an improvement upon existing approaches to estimating the likelihood an individual is autistic (which could allow better prioritization of incoming patients for diagnostic interviews) as well as a neuropsychological profile of strengths and weaknesses to assist clinicians in supporting patients’ specific needs. When one considers the social cost of long wait times for diagnostic interviews in combination with steadily rising rates of diagnoses (particularly among adults), the potential value of a more accurate and informative screening method that is as accessible as internet connection and briefer than a typical movie is clear.

## Data Availability

All data produced in the present study are available upon reasonable request to the authors.

## Footnotes

1 We alternate between person-first and identity-first language to address the varying preferences of the international autism community, as recommended in Buijsman et al. (2023).

2 The ICAR project is an open resource for online cognitive tasks ^43^. We used the ICAR-16 intelligence test, which correlates strongly with full-scale IQ ^44^ and has been shown to be age- and sex-invariant ^45^. We report proportions of correct responses, which we use as covariates in subsequent analyses. Note that the ICAR performance was unknown for four participants.

