## Supplementary Materials for "Finding the Forest in the Trees: Using Machine Learning and Online Cognitive and Perceptual Measures to Predict Adult Autism Diagnosis"

**Description of Features**

In the first [1] of the three studies that comprise the dataset used for this paper, we employed the classic McGurk/MacDonald paradigm to measure a) the ability to integrate audiovisual speech stimuli [2], b) temporal properties for processing multisensory information [3], and c) the ability to rapidly adapt to multisensory asynchronies, known as rapid temporal recalibration [4–6]. The participants viewed a video of an actress mouthing the syllable /ga/. On half the trials, audio corresponded to the video (i.e., congruent trials), whereas on the remaining trials, the audio of the syllable /ba/ was played (i.e., incongruent trials). The onset between the voice and the lip movement was manipulated, and the participants were instructed to make two judgments. First, they reported whether they heard /ba/, /ga/, or /da/. Subsequently, they judged whether the voice was synchronized with the lip movements. For the RF classifier, we used the proportion of /ba/, /ga/, and /da/ responses (i.e., *mcg_ba_response*, *mcg_ga_response*, and *mcg_da_response*), the proportion of synchrony responses (*mcg_sync_response*), and the proportion of synchrony responses for congruent trials – incongruent trials (*mcg_congruency_effect*). Furthermore, Gaussian functions were fitted to the synchrony distributions to estimate the point of subjective simultaneity (i.e., the audiovisual asynchrony whereby the stimuli are most likely to be perceived as synchronous; *mcg_pss*), and the window of perceived synchrony (i.e., the maximum audiovisual asynchrony whereby the stimuli are likely to be perceived as synchronous; *mcg_wps*). Finally, we measured rapid temporal recalibration, which was defined as the PSS following trials in which vision led minus the PSS following trials in which audition led (*mcg_recalibration*).

In the second study [7], we measured the performance on three different emotion recognition tasks. In the basic facial emotion recognition task, participants saw photographs selected from a subset of the KDEF database used in a previous study [8] of actors/actresses making emotional expressions (happy, sad, angry, afraid, disgusted, surprised, and neutral). A photograph would appear along with the 7 word/number pairs, and participants were instructed to press the key corresponding to the emotion they thought the person in the image was experiencing as quickly and accurately as possible. For the RF classifier, we used the mean correct response time (*kdf_rt*), the mean accuracy (*kdf_accuracy*), and the improvement over the course of the experiment in both dependent variables (i.e., the RT and accuracy for the first half of the experiment – the RT and accuracy for the second half of the experiment; *kdf_rt_over_time* and *kdf_accuracy_over_time*).

In the complex facial emotion recognition task, participants saw photographs of the eyes of actors/actresses making various emotional expressions taken from the reading the mind in the eyes test [9]. Four words describing feelings (one of which corresponded to each expression) were displayed, and participants were instructed to press the number key corresponding to the option they felt best described what the person in the image was feeling as quickly and accurately as possible. For the RF classifier, we used the mean correct RT (*rme_rt*), the mean accuracy (*rme_accuracy*), and the mean RT and accuracy over the course of the experiment (*rme_rt_over_time*, and *rme_accuracy_over_time*, respectively).

In the affective prosody recognition task, a trial began with the presentation of a green (go trial) or red (no-go trial) fixation symbol, followed by a semantically neutral Dutch sentence, where affective prosody varied from happy to fearful [10,11]. On go trials, participants were instructed to respond to what emotion the voice expressed on a Likert scale of 1-7 (with 1 being happiest and 7 being most fearful). On no-go trials, participants were asked to withhold their response. For the RF classifier, we used the mean correct response time (*psd_rt*) and the mean response time over the course of the experiment (*psd_rt_over_time*).

We also measured sensitivity to affective prosody. However, we did not include this measure, as we excluded too many participants in our previous study [7].

In the final study [12], we measured performance on four different executive functioning tasks. In the go/no-go task, we measured the ability to inhibit a response [13]. Every trial, participants saw an arrow on the screen and were instructed to press the z- or m-key when the arrow was facing left or right, respectively (i.e., a go trial), or to withhold their response when the arrow was facing up (i.e., a no-go trial). For the RF classifier, we used the mean correct response time on go trials (*gng_rt*), the mean proportion of omission errors (i.e., making no response on go trials; *gng_omission_err*), the mean proportion of commission errors (i.e., making a response on no-go trials; *gng_commission_err*), and the mean proportion of direction errors (i.e., making an incorrect response on go trials; *gng_direction_error*). Furthermore, we used the mean correct RT and overall error over the course of the experiment (*gng_rt_over_time*, and *gng_error_over_time*, respectively). Finally, we measured some inter-trial effects reflecting response inhibition (i.e., the mean correct RT on a go trial when preceded by no-go trial – the mean correct RT on a go trial when preceded by go trial; *gng_rt_inhibition*), motor priming (i.e., the mean correct RT on switch trials – mean correct RT on repetition trials; *gng_motor_priming_rt*, and the mean error rate on switch trials – mean error rate on repetition trials; *gng_motor_priming_error*). A switch trial is defined as a trial in which the arrow on a preceding go trial was facing toward a different direction compared to the arrow on the current go trial. For repetition trials, the arrow was facing the same direction.

In the arrow/gaze cueing task, we measured whether social versus non-social cues affect visual orienting [14]. Trials started with the presentation of a fixation cross, then an arrow (facing to the left or right) or gaze (eyes facing to the left or right) cue appeared. Subsequently, after 100 ms, a red circle (the target) appeared on either the left or right side of the cue, and participants were instructed to press the z- or m-key (respectively) as quickly as possible. On half of the trials, the cue was valid (the eyes or arrow were facing towards the target location), and on the remaining trials, the target was invalid (i.e., faced opposite to the target). As such, the cue was uninformative about the target location (i.e., cue validity was 50%).

In the chessboard task [15], we measured spatial working memory capacity. A trial started with a fixation cross. Subsequently, a 4x4 grid of alternating orange and blue squares separated by thin white borders appeared on the screen. Then, a number of squares would flash serially in lighter versions of their respective colors. The squares that flashed were randomly selected, with the constraints that at least one of each color flashed and that at least one blue square flashed before the last orange one. Participants were instructed to respond by clicking all the orange squares and then all the blue squares, both in the order in which they lit up. The next trial began as soon as the participant clicked as many squares as had flashed (irrespective of whether they were the same ones). The task difficulty (i.e., the number of flashing squares) was adaptive, beginning with a span of three squares flashing. After two consecutive correct/incorrect trials, the span would increase/decrease by one square. For the RF classifier, we used the mean response time (too slow trials excluded; *cbd_rt*), the maximum working memory span (*cbd_max_span*), and the number of errors (too slow trials excluded; *cbd_blue_errors*, *cbd_orange_errors,* and *cbd_color_sequence_errors*).

In the trail making task [16], we measured cognitive flexibility by comparing two different conditions. In the numeric condition (TMT-A), the display consisted of small circles numbered 1-25, and participants were instructed to respond by clicking through the circles as quickly and accurately as possible in numerical order. The alphanumeric condition (TMT-B) included 24 circles with letters (A-L) and numbers (1-12), and participants were instructed to alternate between them in the correct order (e.g., 1-A-2-B-3-C…). On both parts, hovering above a node caused it to turn a light shade of gray. When participants clicked the correct node, it would flash a darker shade of gray for 500 ms, and a line would appear connecting it with the previous one. When they clicked the incorrect node, it would flash red for 500 ms. Participants first completed the numeric condition, followed by the alphanumeric condition, without practice. For the RF classifier, we used the completion time to complete TMT-A and TMT-B (*tmt_compl_time_A* and *tmt_compl_time_B*), and the mean response time over the nodes for TMT-A and TMT-B (outlier trials excluded; *tmt_rt_A* and *tmt_rt_B*). We also used the cognitive flexibility for both the total completion time and the mean response time over the nodes (i.e., part B – part B; *tmt_compl_cogn_flexibility*, and *tmt_cogn_flexibility*). Finally, we used the response time over the course of the experiment for part A and part B (*tmt_rt_A_over_time*, and *tmt_rt_B_over_time*).

**Partial Dependence Plots**

Supplementary Figures 1 and 2 show the partial dependence plots for each significant feature for all participants and the age/gender-matched group, respectively. A partial dependence plot visualizes how a specific feature influences the model's predictions by showing the relationship between that feature and the prediction outcome, while keeping all other features constant. In these plots, the y-axis represents the model's predicted likelihood of an individual being classified as autistic (closer to 1) or not autistic (closer to 0). The x-axis shows the range of values for the specific feature being examined, such as task performance or other variables (like age and gender). This allows to see whether a feature has a straightforward (linear), gradually increasing or decreasing (monotonic), or more complex effect on the prediction. For example, in the case of kdf_rt (reaction time for the basic emotion recognition task), the plot shows that participants with shorter reaction times are slightly more likely to be classified as not autistic, while those with longer reaction times are slightly more likely to be classified as autistic. The most significant changes in classification likelihood occur around a reaction time of 3000 ms. Beyond this point, at both very low and very high reaction times, the likelihood stabilizes, indicating that further changes in reaction time have minimal impact on the prediction.

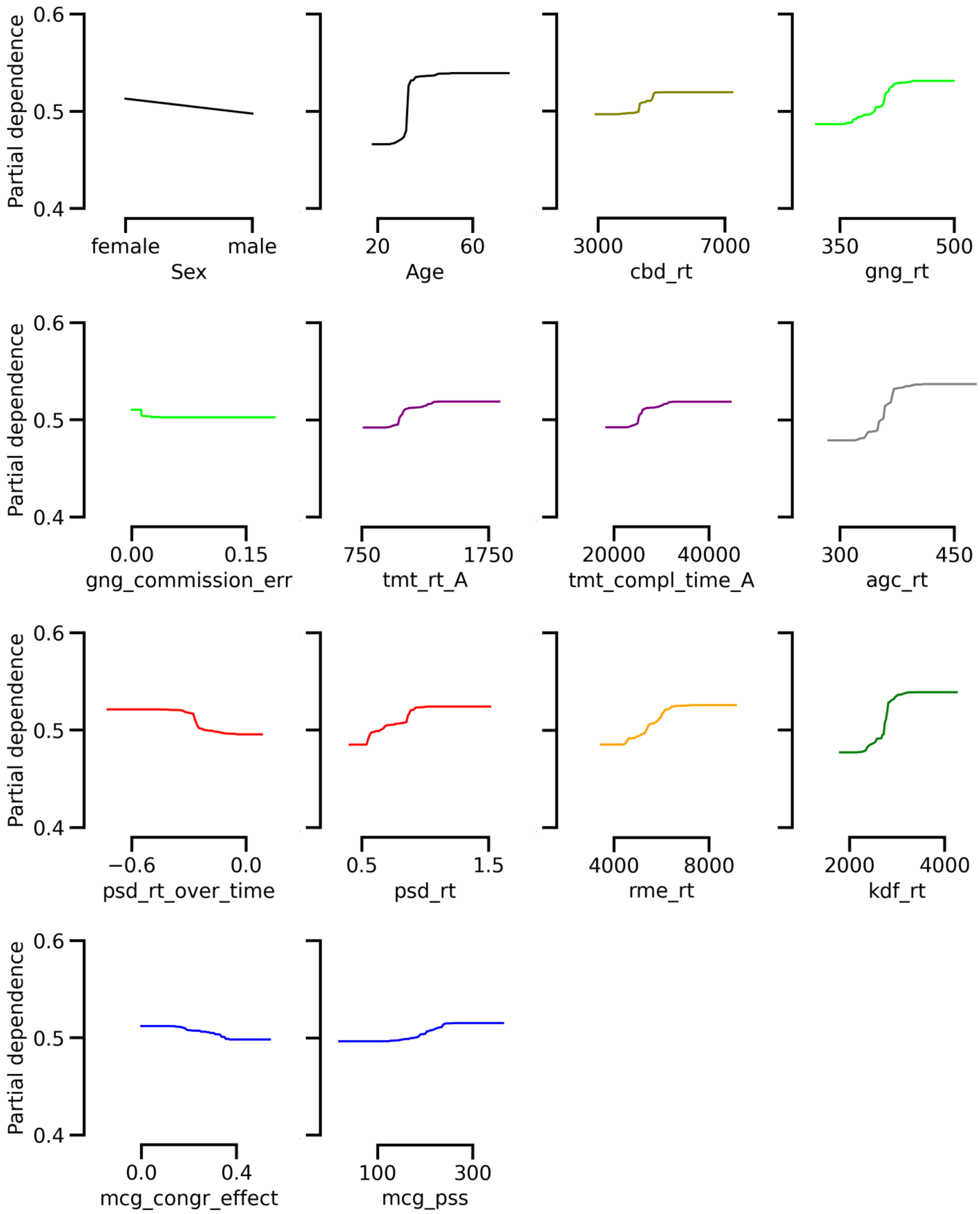


Supplementary Figure 1. Partial dependence for each significant feature in Figure 2A (all participants included). Partial dependence describes the contribution of each feature of the data to the prediction of autism. Note that each color represents a particular task (like in Figure 2A).

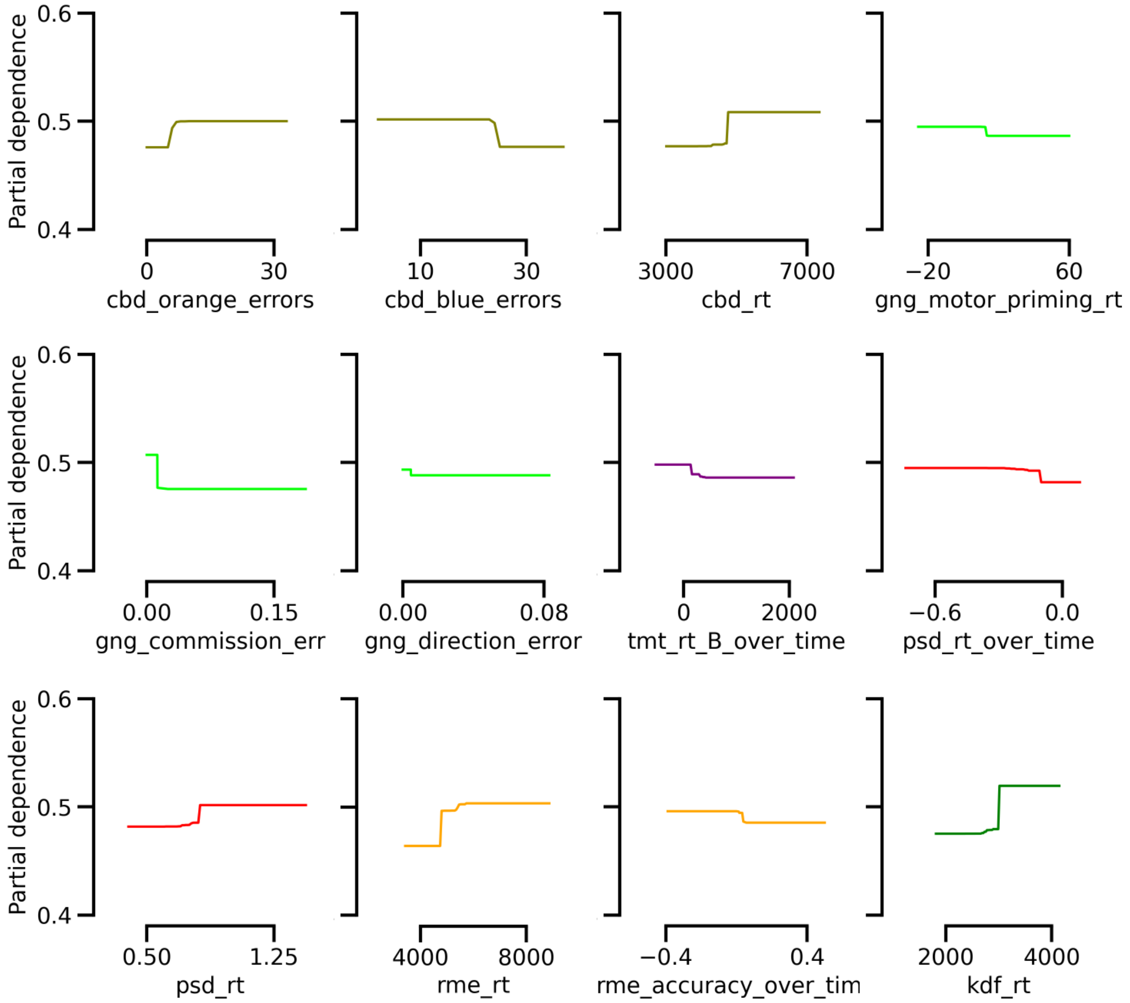


Supplementary Figure 2. Partial dependence for each significant feature in Figure 3A (age- and sex-matched group). Partial dependence describes the contribution of each feature of the data to the prediction of autism. Note that each color represents a particular task (like in Figure 3A).

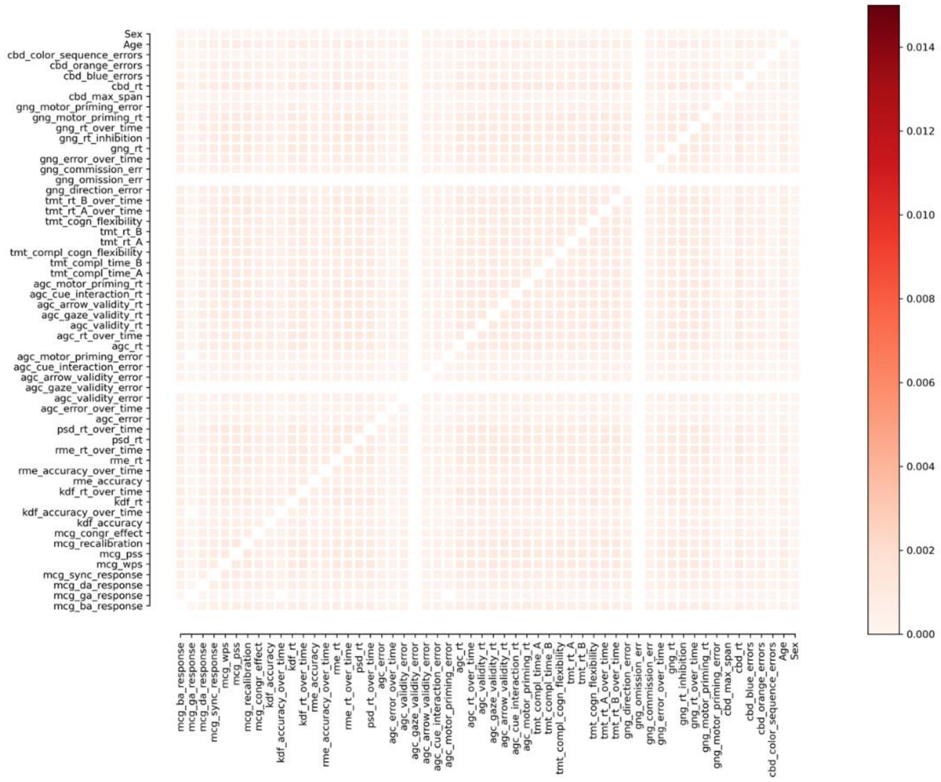


Supplementary Figure 3. Likelihood that features will occur together in the same decision tree when the labels of participants in the full sample were randomized (all values add up to one). Darker blocks indicate features that frequently co-occur and are therefore useful in combination in predicting autism.

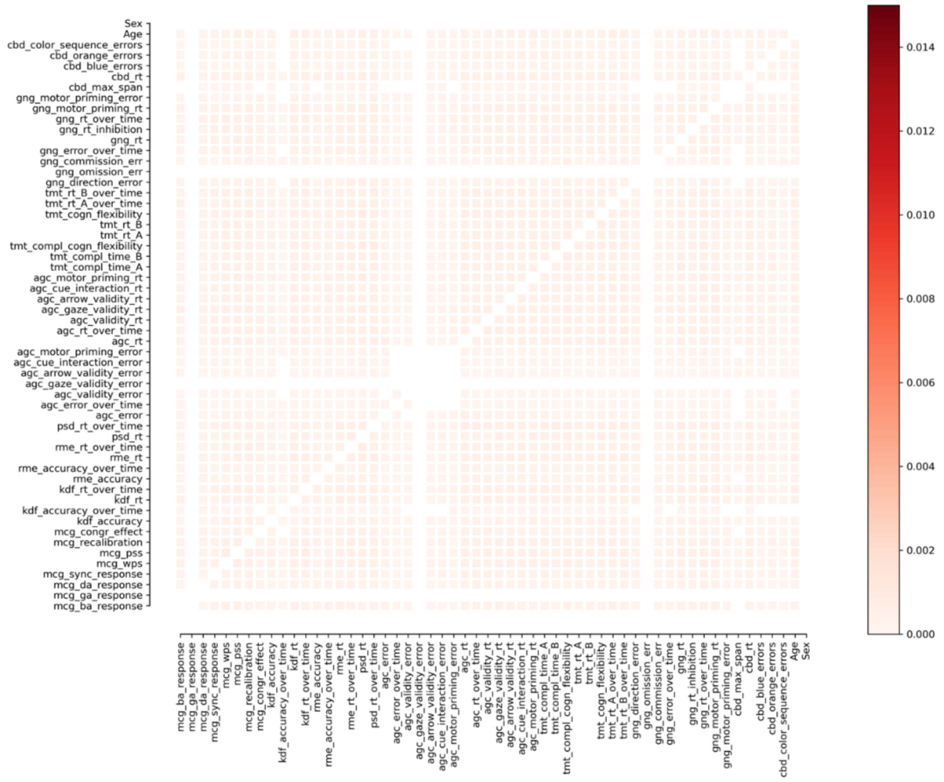


Supplementary Figure 4. Likelihood that features will occur together in the same decision tree when the labels of participants in the age- and sex-matched sample were randomized (all values add up to one). Darker blocks indicate features that frequently co-occur and are therefore useful in combination in predicting autism.

**Model performance and feature importance with AQ-28 as additional feature**

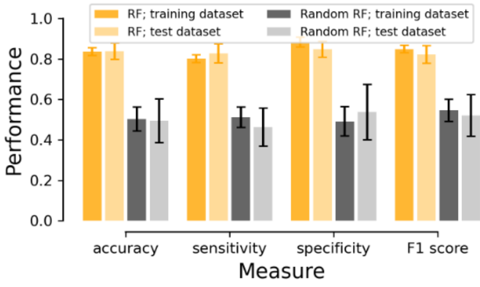


Supplementary Figure 5. The mean performance for the random forest (RF) classifier models with AQ-28 score included as feature for each participant with properly labeled groups (orange bars) and the RF classifier models with the group labels randomized (gray bars) in terms of accuracy, sensitivity, specificity, and F1 score for the age- and gender-matched subgroup. The dark bars signify the performance on the training dataset, whereas the light bars reflect the performance on the test dataset. Error bars represent the standard deviation.

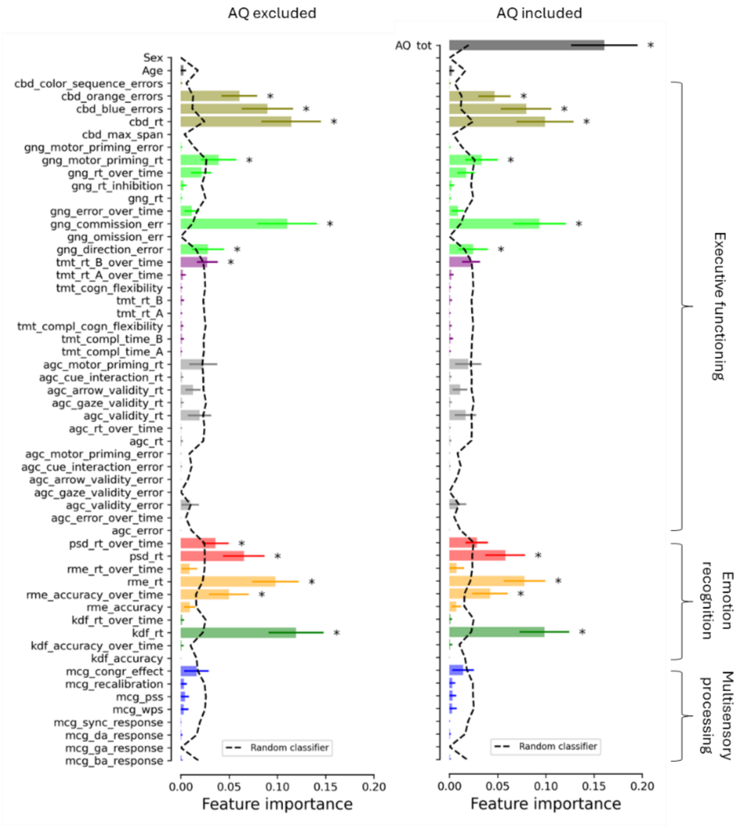


Supplementary Figure 6. Mean feature importance score for each feature in the age- and gender-matched dataset with (same as Figure 3A) vs. without the Autism Quotient 28 included as a feature. Note that each task is identified by a unique color and the first three characters of the corresponding feature names (mentioned in the Method section). Here, important features are indicated by an asterisk, indicating that the feature score for the RF model was significantly ($p < .05$, Bonferroni corrected) larger than the feature score for the random RF model (see the dashed line). Error bars represent the standard deviation.
